# Haloperidol inhibits inflammasome activation via LAMTOR1 and reduces the risk of arthritides

**DOI:** 10.1101/2023.12.06.23299609

**Authors:** Vidya L. Ambati, Praveen Yerramothu, Ranjith Konduri, Joseph Nguyen, Bradley D. Gelfand, E. Will Taylor, Brian C. Werner, Shao-bin Wang

## Abstract

Gout is the most prevalent form of inflammatory arthritis in the world. Although multiple treatments exist, many patients are poorly responsive. Here we report, using a health insurance database analysis, that use of the anti-psychotic haloperidol is associated with a reduced risk of incident gout. Haloperidol inhibits ASC speck formation, caspase-1 activation, and release of IL-1β and IL-6, suggesting that it inhibits NLRP3 inflammasome activation and downstream cytokine responses. We also identified LAMTOR1 as a novel binding partner for haloperidol and demonstrate that haloperidol inhibits the aggregation of LAMTOR1 and NLRP3. Since NLRP3 inflammasome activation has been implicated in gout, these data provide a foundation for exploring haloperidol as a potential therapy.

## Introduction

Gout is the most common form of inflammatory arthritis and is estimated to affect 2 to 3% of the world’s population (1). Over the last three decades, the incidence of gout has roughly doubled in most of the world (2). Typically, gout is characterized by episodic acute inflammatory events or flares (3). A common feature of gout is the deposition of monosodium urate (MSU) crystals in and around joints (4). A principal therapy for gout is the reduction of serum urate levels to enable dissolution of deposited crystals (5, 6). Other first-line therapies include non-steroidal anti-inflammatory drugs, colchicine, and corticosteroids to limit the inflammation and pain that accompany gout flares. Unfortunately, many patients poorly tolerate these therapies or respond inadequately (7). Urate crystals activate the NLRP3 inflammasome (8) and inhibitors of interleukin-1β (IL-1β), a cytokine released by inflammasome activation (9), are also part of the therapeutic armamentarium against gout (10, 11). However, IL-1β inhibitors carry a high economic cost as well as a serious adverse event profile.

Although several drugs are approved for the management of gout, most have significant potential side effects. Therefore, there is a need to explore alternate therapeutic strategies. One such approach is to identify existing FDA-approved drugs that could be repurposed as treatments for new indications (12). Recently, we reported that exposure to haloperidol, an FDA-approved drug for the treatment of schizophrenia or Tourette’s disorder, was associated with reduced risk of incident rheumatoid arthritis (13). Since inflammasome activation plays a critical role in both rheumatoid and gouty arthritis (8, 14), we also investigated whether haloperidol affects this inflammatory process.

## Methods and Materials

### Health Insurance Claims Databases Analyses

Health insurance database information contains de-identified data that are Health Insurance Portability and Accountability Act (HIPAA)-compliant and were deemed by the University of Virginia Institutional Review Board (IRB) as exempt from IRB approval requirements. The retrospective study used claims data from the PearlDiver Mariner database, which contains data on health care claims and medication usage for persons in provider networks over the time period 2010 to 2022.

Patients were included in the analysis if they had continuous enrollment in the plan for at least 6 months, were at least 18 years of age at baseline, and were confirmed to have schizophrenia or Tourette’s disorder (diagnosed on at least 2 separate occasions). Individuals with pre-existing gout prior to diagnosis of schizophrenia or Tourette’s disorder were excluded. Disease claims were identified by International Classification of Diseases (ICD)-9-CM and ICD-10-CM codes. Exposure to haloperidol or other anti-psychotics – the independent variable – was determined by whether patients filled pharmacy prescriptions for generic or brand versions, as identified by National Drug Codes. Time from diagnosis of schizophrenia or Tourette’s to initial diagnosis of gout was the dependent variable.

Analyses were performed using R Studio, Version 2023.06.0+421 (the R project). To analyze the risk of gout between haloperidol users and haloperidol non-users (those who used other anti-psychotics), an adjusted Cox proportional hazards regression analysis was performed, and the hazard ratio was analyzed. The adjusted model included these confounding variables: age, sex, smoking, body mass index, and Charlson Comorbidity Index, and year of plan entry. The haloperidol-exposed and haloperidol-unexposed groups were also matched for these variables using greedy nearest neighbor propensity score matching using the R package MatchIt. Cox models were analyzed by chi square test. Patients were censored when they developed gout, left the plan, or switched to another class of anti-psychotic. Statistical tests were 2-sided. P values < 0.05 were considered statistically significant.

### Cell culture studies

All cell culture experiments were compliant with University of Virginia Institutional Biosafety Committee regulations. Human THP-1 cells (American Type Culture Collection) were differentiated into macrophages with 0.5 μM phorbol 12-myristate 13-acetate (PMA, Sigma Aldrich) and cultured in RMPI-1640 media (Thermo Fisher) supplemented with 10% fetal bovine serum (FBS) and 1% penicillin–streptomycin. Human fibroblasts (Cell Applications) were cultured in HFLS medium (Cell Applications). Cells were maintained at 37 °C in a 5% CO_2_ environment.

### ASC speck imaging

THP-1-derived macrophages were seeded on chambered coverslips (30,000 cells/well) for 12 h and pretreated with haloperidol (1 μM, Sigma-Aldrich) or 0.1% DMSO (control) for 2 h. Cells were treated with LPS (125 ng/mL) for 4 h and ATP (5 mM) for 15 min. Eight well chamber plates (Grace Bio Labs) were fixed with 2% paraformaldehyde for 15 min at room temperature, washed with PBS, permeabilized, blocked with blocking buffer (PBS, 0.1% TX-100, 5% normal donkey serum; 1 h at 4 °C), incubated with rabbit anti-human ASC antibody (AdipoGen, 1:200) with blocking buffer, followed by incubation with goat anti-rabbit-488 (Thermo Fisher, 1:500). DAPI-stained slides were mounted using Fluoromount-G (Southern Biotech) and imaged by confocal microscopy (Nikon A1R). The number of ASC specks per 0.09 mm^2^ field was quantified. Means were compared using two-tailed Student t test.

### Caspase-1 western blotting

THP-1 cells were treated with LPS and ATP and pre-treated with haloperidol (0.1–1 μM) for 1 h. Proteins from the cell-free supernatant were precipitated by adding sodium deoxycholate (0.15% final), followed by adding TCA (7.2% final) and incubating on ice overnight. Samples were spun down at 12000g for 30 min and pellets were washed 2 times with ice-cold acetone. Precipitated proteins solubilized in 4X LDS Buffer with 2-mercaptoethanol were resolved by SDS-PAGE on Novex® Tris-Glycine Gels (Invitrogen) and transferred onto low fluorescence PVDF membranes (Bio-Rad). The transferred membranes were blocked with LI-COR block for 1 h at room temperature and then incubated with anti-human caspase-1 antibody (AdipoGen, 1:1000) at 4 °C overnight. The immunoreactive bands were labeled using species-specific secondary antibodies conjugated with IRDye®. Blot images were captured using an Odyssey® imaging system.

### IL-1β and IL-6 ELISA

THP-1 cells and fibroblasts were treated with LPS and ATP and treated with haloperidol (0.01– 100 µM) as above. Secreted IL-β and IL-6 in the conditioned cell culture media were detected using ELISA kits (R&D Systems) according to the manufacturer’s instructions. Means were compared using two-tailed Student t test or one-way ANOVA with post hoc Tukey’s test.

### Biotin-haloperidol synthesis

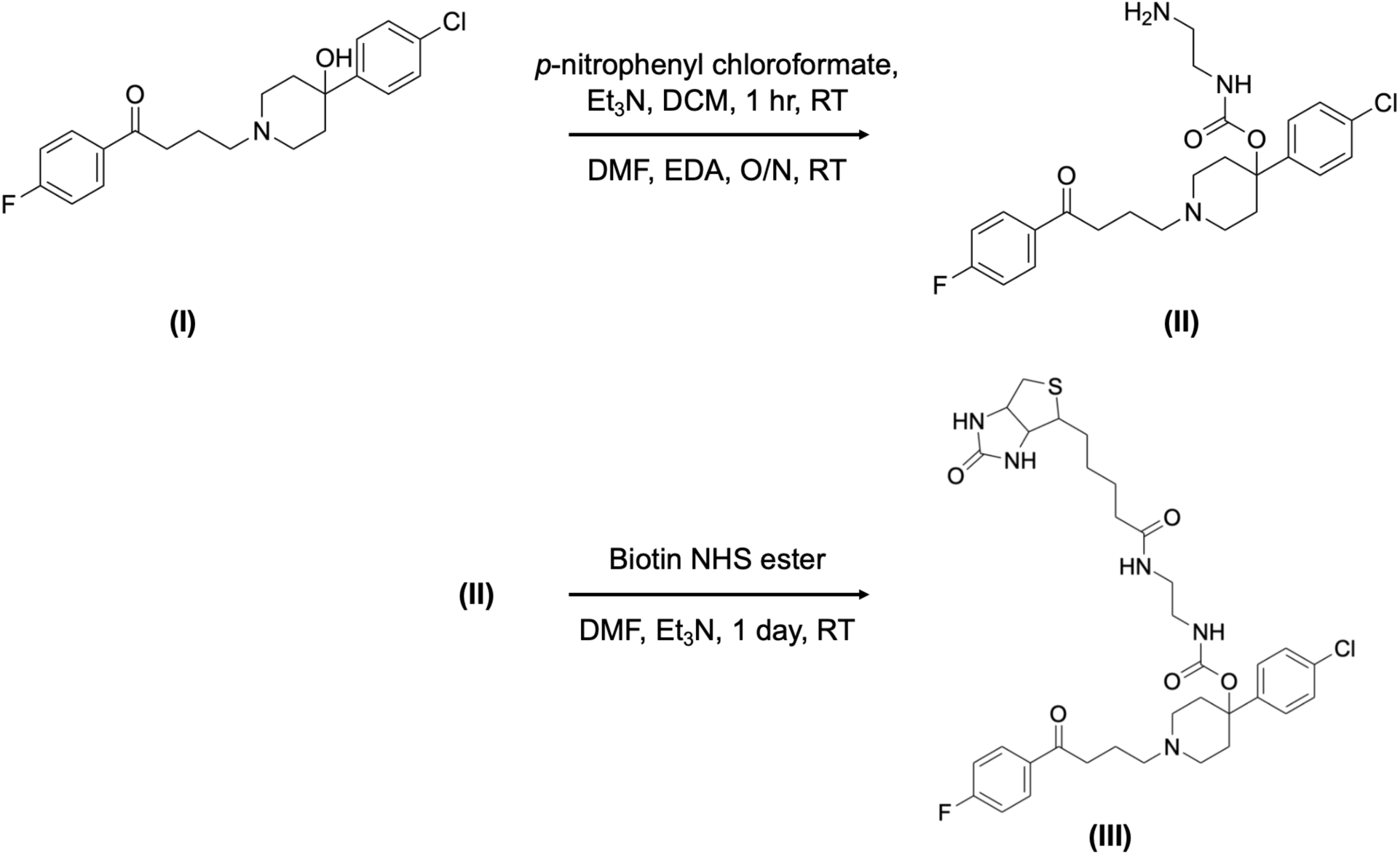

Haloperidol (**I**) (150 mg, 0.4 mmol) was dissolved in anhydrous dichloromethane (DCM, 10 mL). To this solution, *p*-nitrophenyl chloroformate (160 mg, 0.8 mmol) and triethylamine (Et_3_N, 139 μL, 1.0 mmol) were added. The reaction was stirred for 1 hour at RT and then added dropwise to a solution of ethylenediamine (EDA, 267 μL, 4 mmol) in dimethylformamide (DMF, 20 mL). The reaction was stirred overnight. The haloperidol-aminoethyl carbamate (haloperidol-AEC) (**II**) product was purified by washing with 0.1 M sodium hydrocarbonate (3 × 200 mL), followed by chromatography on a SiO_2_ column (30 × 2.5 cm), eluted with a chloroform-methanol mixture (90 : 10 to only methanol) and determined by TLC analysis.

### Streptavidin pulldown

To detect binding partners of haloperidol, LPS-primed wild-type mouse bone marrow-derived macrophages (BMDMs) were lysed with NP-40 lysis buffer (50 mM Tris-HCl pH 7.4, 150 mM NaCl, 1% NP-40, 5 mM EDTA and 0.1% Triton-X 100) on wet ice, and centrifuged at 12,000 rpm for 15 minutes at 4°C. The lysate supernatant was then pre-cleared by streptavidin magnetic beads (88816, Thermo Fisher) to remove nonspecific binding. Pre-cleared lysates were incubated with free haloperidol and biotinylated haloperidol, as indicated, for 1 hour in an ice bath. Then, these samples were incubated with pre-activated streptavidin magnetic beads overnight at 4°C with rotation. On the next day, the beads were washed with lysis buffer three times, and then boiled with SDS sample buffer (LC2676, Thermo Fisher) for mass spectrometry analysis.

For in vitro binding and competition assays, recombinant His-PDCD1-tagged human LAMTOR1 protein (TP762012, OriGene) was precleared using streptavidin magnetic beads (88816, Thermo Fisher) to remove nonspecific binding and then incubated with biotinylated haloperidol or free (unbiotinylated) haloperidol for 1 h on ice. These samples were then incubated with preactivated streptavidin magnetic beads overnight at 4 °C with rotation. On the next day, the beads were washed with NP-40 lysis buffer three times and then boiled with sodium dodecyl sulfate (SDS) sample buffer (LC2676, Thermo Fisher) for further analysis.

### Untargeted Protein Identification and Label-free Quantification via Tandem Mass Spectrometry

Bead-bound proteins were released by treatment with 100 mM ammonium bicarbonate, pH 7.5. Samples were reduced with 5 mM dithiothreitol for 90 min at room temperature, and alkylated with 10 mM iodoacetamide for 45 min at room temperature, protected from light. Modified porcine trypsin protease (Promega #V5113) was added at a ratio of 1:20 w/w enzyme:protein at 37°C and incubated overnight, and the digestion was quenched with formic acid. Peptides were then desalted by reverse-phase chromatography using Pierce peptide desalting spin columns (Thermo Fisher #89852), dried completely, and resuspended in Solvent A (0.1% formic acid in Fisher Optima LC/MS grade water). Samples were quantified by A_280_ absorbance and total injection amount was normalized.

Samples were subjected to mass analysis using a Thermo Scientific Ultimate 3000 RSLCnano ultra-high performance liquid chromatography (UPLC) system coupled to a high-resolution Thermo Scientific Eclipse Tribrid Orbitrap mass spectrometer. These analyses were performed at the University of Connecticut Proteomics & Metabolomics Facility. Each sample was injected onto a nanoEase M/Z Peptide BEH C18 column (1.7 μm, 75 μm x 250 mm, Waters Corporation) and separated by reversed-phase UPLC using a gradient of 4-90% Solvent B (0.1% formic acid in Fisher Optima LC/MS grade acetonitrile) over a 60-min gradient at 300 nL/min flow, followed by a 10-min wash and 20-min column re-equilibration. Peptides were eluted directly into the Eclipse using positive mode nanoflow electrospray ionization with source spray voltage set to 2200 V. MS1 scans were acquired at 120,000 resolution with AGC target set to Standard, maximum injection time set to Auto, RF lens set at 30%, and scan range set for 300-1800 m/z. Data-dependent MS2 scans were collected for ions in charge states 2-8 at a minimum intensity of 5.0e4. Scans were acquired in the Orbitrap at 15,000 resolution with AGC target set to Standard, maximum injection time set to Dynamic, isolation window set to 1.6 m/z, for a cycle time of 3 sec. Fragmentation was performed using HCD at 30% energy. Dynamic exclusion was enabled after 1 observation, for 30 sec, with a tolerance window of 20 ppm total.

Peptides were identified using MaxQuant (v1.6.10.43) and its embedded Andromeda search engine and quantified by label-free quantification (15). The raw data were searched against both the complete UniProt *Mus Musculus* reference proteome (identifier UP000000589, accessed 12 Jan 2022) and the MaxQuant contaminants database. Variable modifications allowed oxidation of Met, acetylation of protein N-termini, deamidation of Asn/Gln, and peptide N-terminal Gln to pyroGlu conversion. Carbamidomethylation of Cys was set as a fixed modification. Protease specificity was set to trypsin/P with a maximum of 2 missed cleavages. All results were filtered to a 1% false discovery rate at the peptide and protein levels using the target-decoy approach; all other parameters were kept at default values. MaxQuant output files were imported into Scaffold (Proteome Software, Inc.) for data visualization and subsequent analyses.

## Results

### Haloperidol associated with reduced risk of gout

We evaluated haloperidol, in the PearlDiver Mariner (151 million people from 2010 to 2022) database, by comparing patients diagnosed with schizophrenia or Tourette’s disorder (the population at risk) on the basis of use of haloperidol versus use of other anti-psychotic drugs. We performed a retrospective, longitudinal cohort analysis using a Cox proportional hazards regression analyses to estimate the hazard of gout in relation to haloperidol use. Since individuals in this database were not randomly assigned to haloperidol treatment, we performed propensity score matching, a causal inference approach (16), to assemble cohorts with similar baseline characteristics, thereby reducing possible bias in estimating treatment effects. Additionally, to control for any residual covariate imbalance, we adjusted for confounders associated with gout: age, sex, smoking, and body mass index, Charlson comorbidity index (a measure of overall health), and year of entry into the database. The adjusted Cox proportional hazards regression model in the propensity-score-matched populations showed a protective association of haloperidol use against incident gout (adjusted hazard ratio = 0.762; 95% CI, 0.713, 0.813; P < 0.001) (**Table 1**).

**Table 1.**

|  | **HR (95% CI)** |
| --- | --- |
| **Haloperidol** | **0.762 (0.713–0.813)** |
| Age | 1.045 (1.043–1.048) |
| Sex (Male) | 2.247 (2.095–2.410) |
| Smoker | 1.104 (1.032–1.181) |
| BMI 30–40 | 1.398 (1.300–1.502) |
| CCI | 1.170 (1.152–1.189) |

HR, hazard ratio. CI, confidence interval. BMI, body mass index. CCI, Charlson comorbidity index

### Haloperidol inhibits inflammasome activation

Since haloperidol use was associated with reduced risk of rheumatoid arthritis (13) and gout, we sought to understand the mechanisms by which haloperidol could confer such protection. Inflammasome activation is considered a key driver of rheumatoid arthritis and gout since patients with rheumatoid arthritis and gout exhibit high levels of the inflammasome constituents NLRP3, ASC, or Caspase-1 (14, 17). In addition, lipopolysaccharide (LPS) and adenosine triphosphate (ATP), which activate the inflammasome (18), are also elevated in patients with rheumatoid arthritis (19–21). Similarly, uric acid, which also activates the inflammasome (8), is elevated in patients with gout (4).

First, we tested whether haloperidol could inhibit inflammasome assembly by monitoring ASC speck formation (22) in THP-1-derived macrophages using immunofluorescence staining. Haloperidol inhibited LPS and ATP-induced ASC speck formation (**Figure 1**) (P = 0.007), indicating that haloperidol blocked inflammasome assembly.

**Figure 1.**
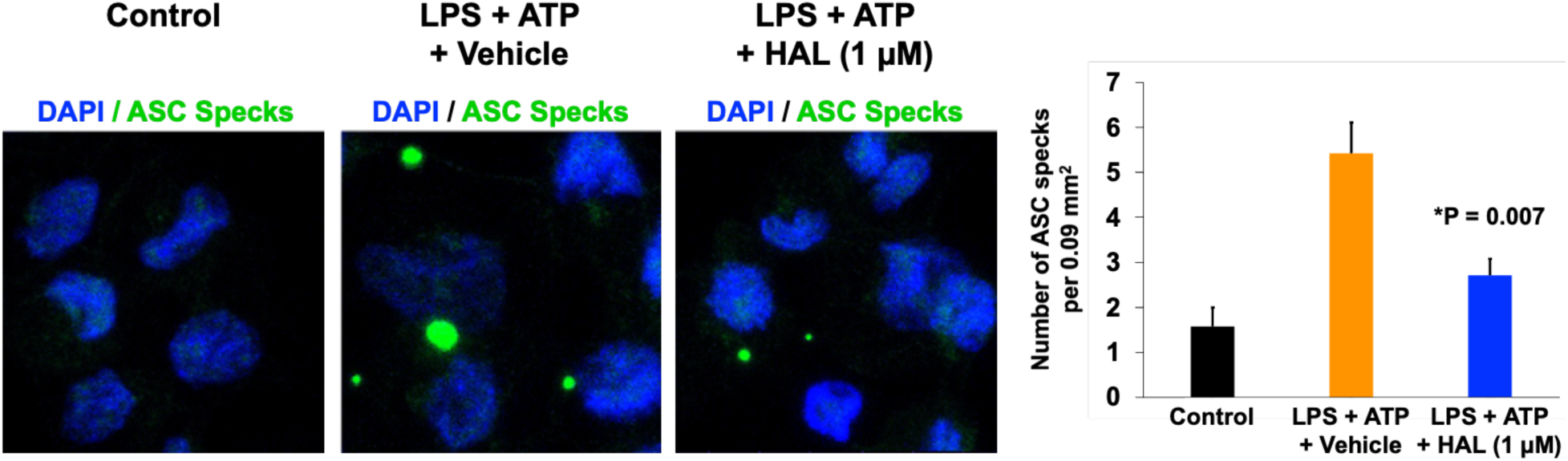
Representative immunofluorescent images show LPS+ATP induces ASC specks (green circular aggregates) in human THP-1 cells, and haloperidol (HAL) reduces speck formation. Cell nuclei stained blue by DAPI. Bar graph of mean and SEM. N = 7 per group. *P=0.007 (LPS+ATP+HAL compared to LPS+ATP+Vehicle), two-tailed Student t test.

Next, we monitored inflammasome activation by assessing proteolytic cleavage of inactive pro-caspase-1 (p45) into active caspase-1 fragments (p33) (18) in THP-1-derived macrophages, using western blotting. Haloperidol robustly inhibited LPS and ATP-induced caspase-1 cleavage, confirming that haloperidol inhibits inflammasome activation (**Figure 2**).

**Figure 2.**
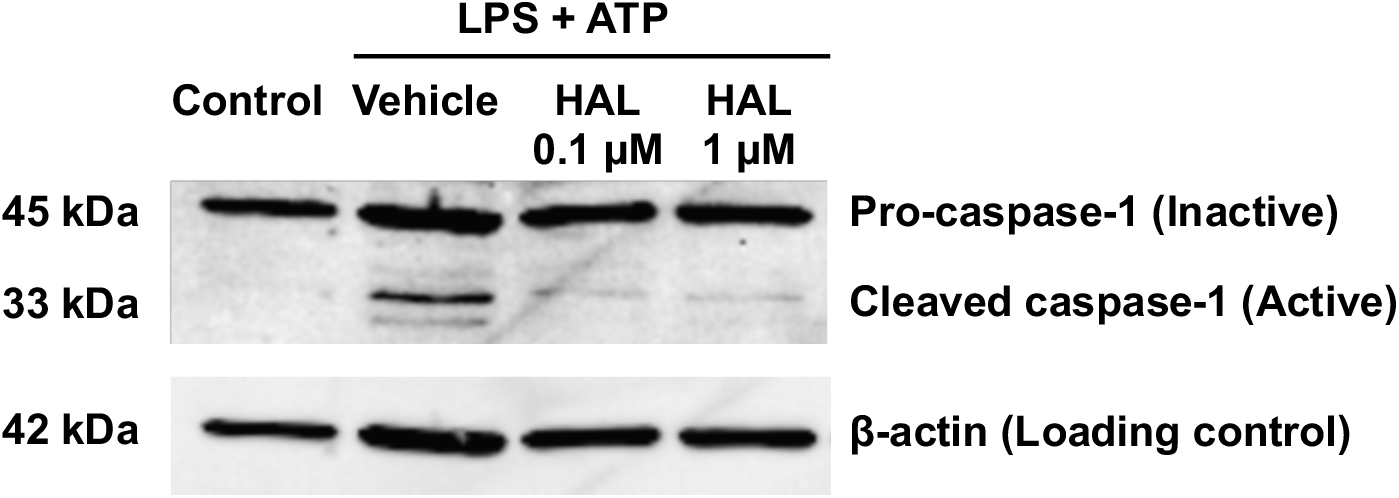
Representative western blot images show LPS+ATP induces cleavage of pro-caspase-1 (45 kDa) into active caspase-1 (33 kDa) in human THP-1 macrophages, and haloperidol (HAL) reduces caspase-1 activation induced by LPS+ATP. β-actin loading control shows protein loading per lane. N = 3 per group.

Finally, we used ELISA to quantify levels of interleukin (IL)-1β, a cardinal inflammasome output, and IL-6, which lies mechanistically downstream of IL-1β, both of which are elevated in rheumatoid arthritis patients (23) and gout (24, 25). In rheumatoid arthritis, the principal cellular source of IL-1β is macrophages and that of IL-6 is synovial (joint) fibroblasts (26). We found that IL-1β release in macrophages was robustly induced by LPS and ATP stimulation, and it was reduced by exposure to haloperidol in a dose-dependent manner (P < 0.05) (**Figure 3**). The potential downstream impact of inflammasome activation was assessed in human synovial fibroblasts by measuring IL-6 secretion. IL-6 release was robustly induced by LPS and ATP stimulation, and it was reduced by haloperidol in a dose-dependent manner (**Figure 3**).

**Figure 3.**
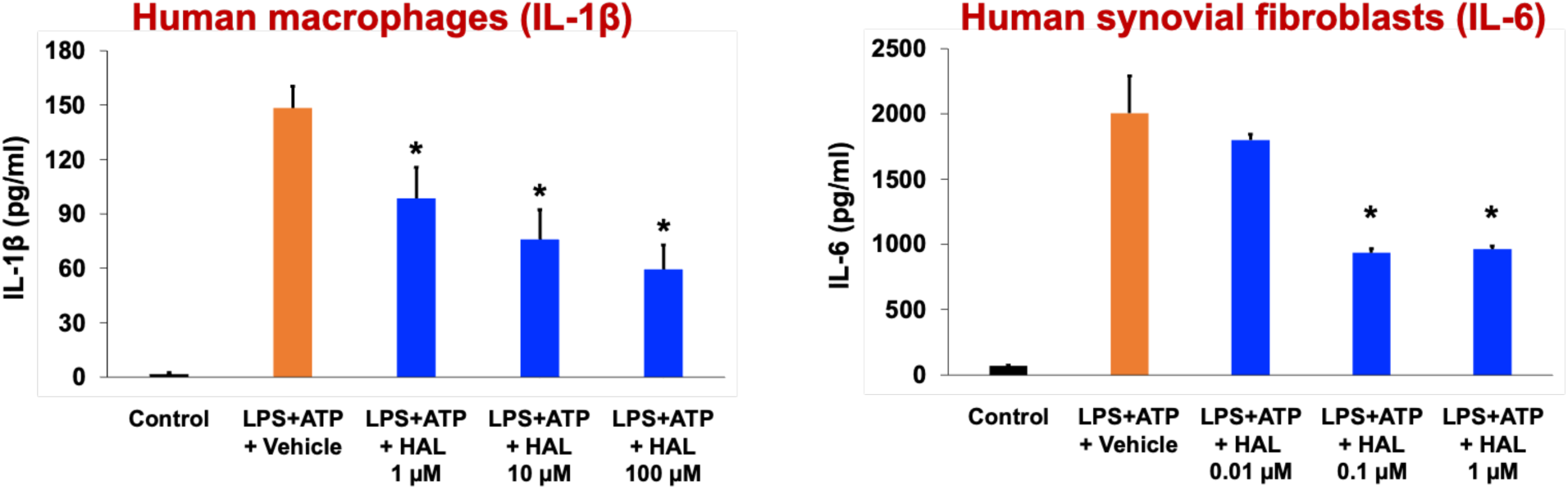
ELISAs of secreted IL-1β from human THP-1 macrophages **(left**) and IL-6 from human synovial fibroblasts **(right**), showing LPS+ATP induces secretion of these cytokines, and haloperidol (HAL) reduces secretion induced by LPS+ATP. N = 3 per group. Data show mean and SEM. *P<0.05 (LPS+ATP+HAL compared to LPS+ATP+Vehicle), two-tailed Student t test.

Similarly, IL-1β release in macrophages was robustly induced by MSU stimulation, and it too was reduced by exposure to haloperidol in a dose-dependent manner (P < 0.05) (**Figure 4**).

**Figure 4.**
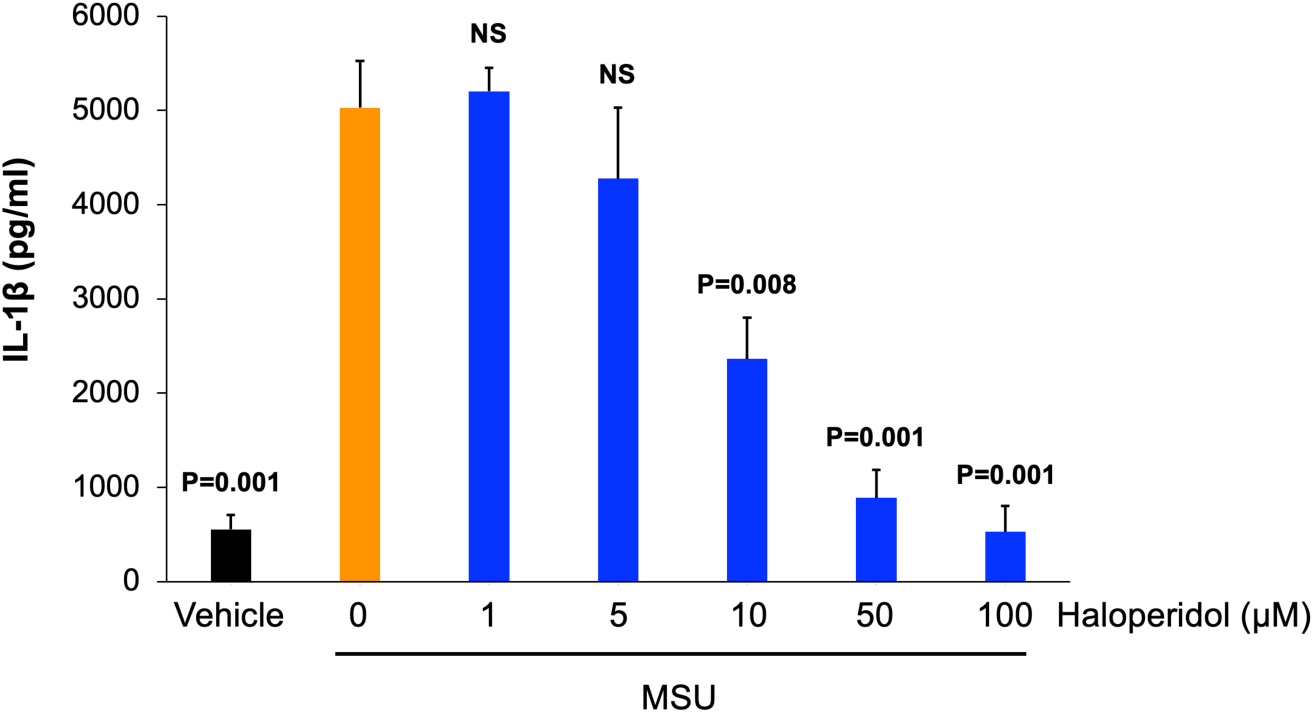
ELISA of secreted IL-1β from human THP-1 macrophages, showing MSU-induced secretion of IL-1β is reduced by haloperidol in a dose-dependent manner. N = 3 per group. Data show mean and SEM. P values (compared to MSU+Vehicle), one-way ANOVA with post-hoc Tukey’s test. NS, not significant.

These data demonstrate that haloperidol inhibits inflammasome activation and subsequent inflammatory pathways.

### Haloperidol interacts with LAMTOR1

Haloperidol is presumed to exert its anti-psychotic effects via its antagonism of the D2 dopamine receptor. Given this novel inflammasome-inhibitory activity we identified, we sought to determine whether haloperidol might have another receptor. Therefore, we performed mass spectrometry proteomics using a pull-down strategy with a biotin-conjugated haloperidol affinity probe. We synthesized this probe using an intermediate aminoethylcarbamate derivative of haloperidol, which provided a handle upon which a biotin linker could be attached. We then incubated LPS-primed wild-type mouse BMDM lysates with biotinylated haloperidol and performed a streptavidin pull down. To identify specific binding partners, we performed a competition control with free (unconjugated) haloperidol. We also used a biotin alone pull down as a negative control. Using tandem mass spectrometry, we found the top specific hit to be the lysosomal protein LAMTOR1 (**Figure 5A**).

**Figure 5.**
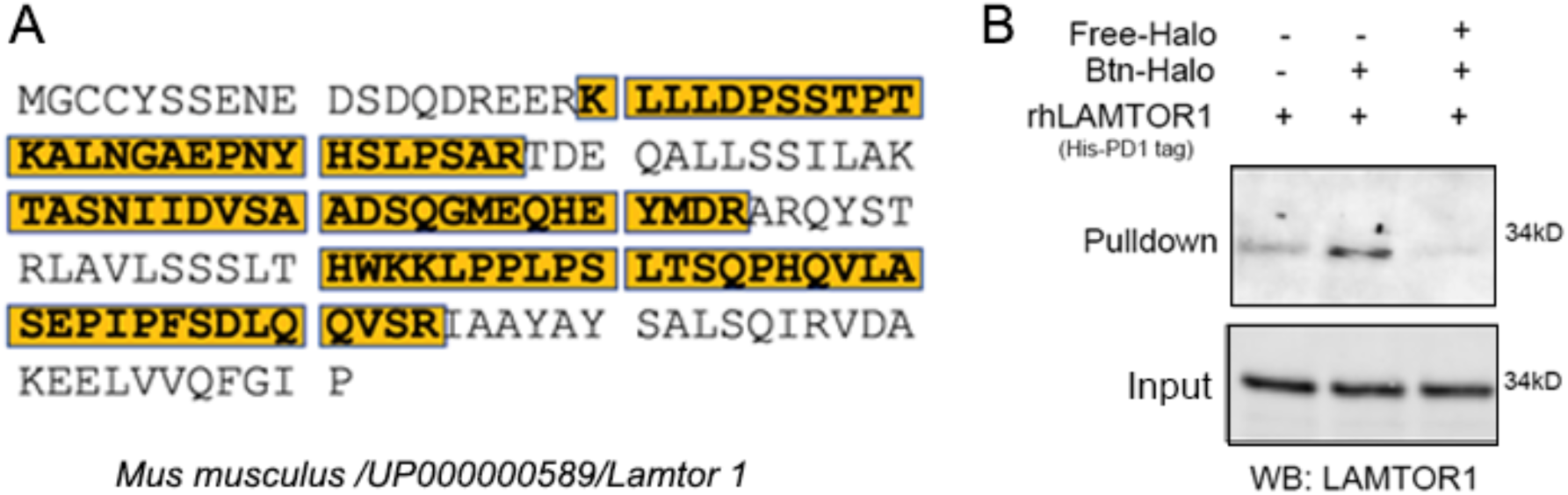
**(A).** Amino acid sequence of mouse LAMTOR1 with amino acid residues identified in mass spectrometry highlighted. (**B**). In vitro interaction of biotinylated haloperidol (Btn-Halo) with recombinant His-PDCD1-LAMTOR1 protein analyzed by streptavidin pull down shows interaction between biotinylated haloperidol and LAMTOR1 that was competed by free haloperidol (Free-Halo). The input and pulldown fractions were immunoblotted for LAMTOR1.

Multiple peptides corresponding to LAMTOR1, collectively comprising 52% coverage of the protein, were identified in the pull down with biotinylated haloperidol. In contrast, no peptides corresponding to LAMTOR1 were identified in the pull downs with biotin alone or when biotin-haloperidol was incubated along with an excess of free haloperidol as a competitor (**Table 2**).

**Table 2.**
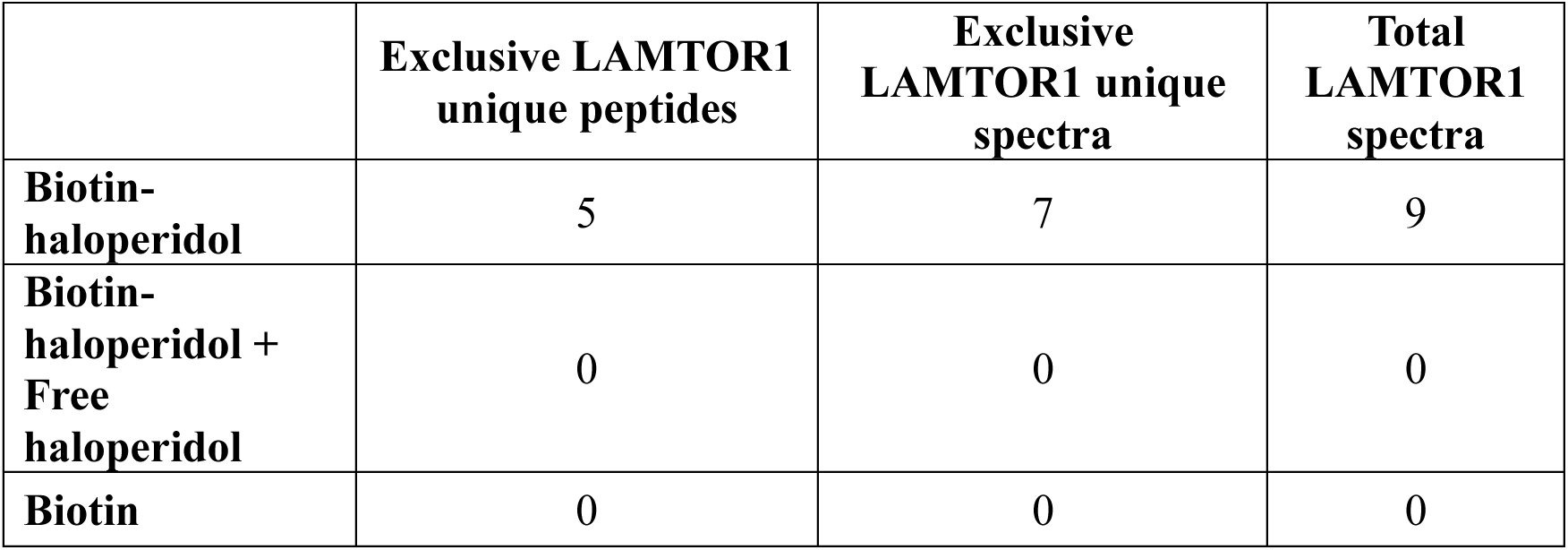

|  | Exclusive LAMTOR1<br>unique peptides | Exclusive<br>LAMTOR1 unique<br>spectra | Total<br>LAMTOR1<br>spectra |
| --- | --- | --- | --- |
| **Biotin-haloperidol** | 5 | 7 | 9 |
| **Biotin-haloperidol + Free haloperidol** | 0 | 0 | 0 |
| **Biotin** | 0 | 0 | 0 |

Next, to confirm the mass spectrometry detection of an interaction of haloperidol with LAMTOR1, we performed an in vitro binding assay incubating recombinant His-PDCD1 tagged human LAMTOR1 protein and biotinylated haloperidol. We were able to detect an interaction between LAMTOR1 and biotinylated haloperidol that was competed by free (unconjugated) haloperidol (**Figure 5B**). Together, these data suggest that haloperidol specifically binds LAMTOR1.

### Computational model of haloperidol-LAMTOR1 interaction

To better understand the interaction between haloperidol and LAMTOR1, we performed a protein-ligand docking study using the SeamDock web server (28). LAMTOR1 exists in a complex with LAMTOR2, LAMTOR3, LAMTOR4, and LAMTOR5 in a structure known as the Ragulator complex (29). Therefore, we submitted the axial conformation of haloperidol and the 5YK3 PDB structure of the Ragulator complex (29) to the SeamDock docking engine. An energetically favorable docking complex was identified with a Gibbs free energy of –8.1 kcal/mol (**Figure 6**). This high-affinity complex corresponds to a dissociation constant (K_d_) of ∼2 μM at 37 °C, which is compatible with the observed IC_50_ of ∼8 μM for haloperidol in inhibiting IL-1β release in cell culture models.

**Figure 6.**
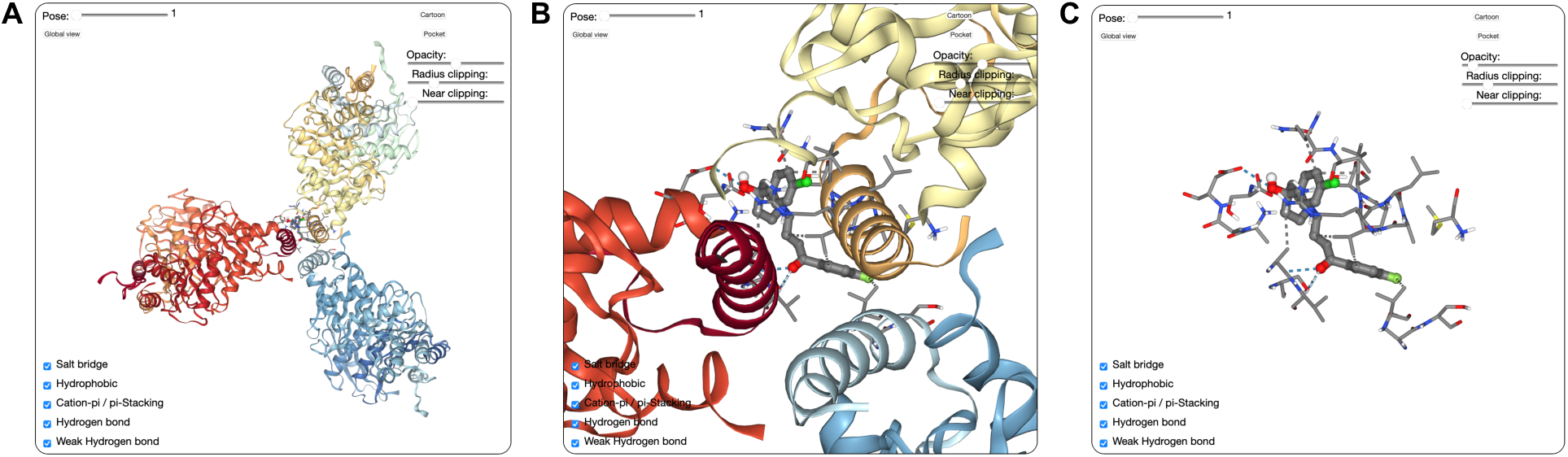
In silico binding analysis of haloperidol with the Ragulator complex. The highest ranked docking complex, whose binding energy was calculated as ΔG = –8.1 kcal/mol, is shown.

In this model, haloperidol binds to 5 amino acids of LAMTOR1 and 2 amino acids of LAMTOR3. This binding is sustained by 6 hydrophobic bonds and 6 hydrogen bonds. Hydrophobic bonds were identified between haloperidol and LAMTOR1 (amino acids 93, 94, 95) and LAMTOR3 (amino acid 119). Hydrogen bonds were identified between haloperidol and LAMTOR1 (amino acids 93, 94, 97, 98) and LAMTOR3 (amino acid 3). Another docking complex in an alternate pose with a Gibbs free energy of –7.9 kcal/mol in a very similar pose was also identified with interactions in similar regions (**Figure 7**).

**Figure 7.**
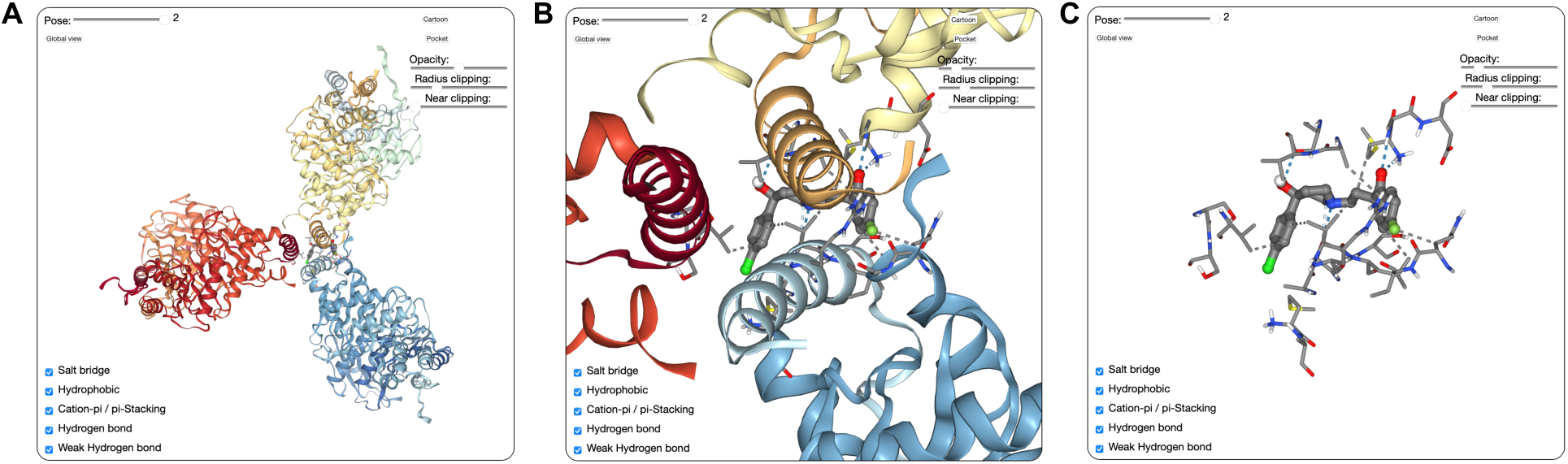
In silico binding analysis of haloperidol with the Ragulator complex. An alternate docked pose, whose binding energy was calculated as ΔG = –7.9 kcal/mol, is shown.

### Haloperidol disrupts with LAMTOR1-NLRP3 aggregation

We found the interaction between haloperidol and LAMTOR1 interesting because LAMTOR1 was recently identified to be a binding partner for NLRP3 that was also critical for inflammasome activation (27). We found that LPS+ATP induced peri-nuclear aggregation of NLRP3 and LAMTOR1 in human THP-1 cells, confirming the recent report (27). Interestingly, treatment with haloperidol markedly reduced the aggregation of NLRP3 and LAMTOR1 (**Figure 8**). These data are compatible with haloperidol binding LAMTOR1 and disrupting its association with NLRP3, thereby reducing inflammasome activation.

**Figure 8.**
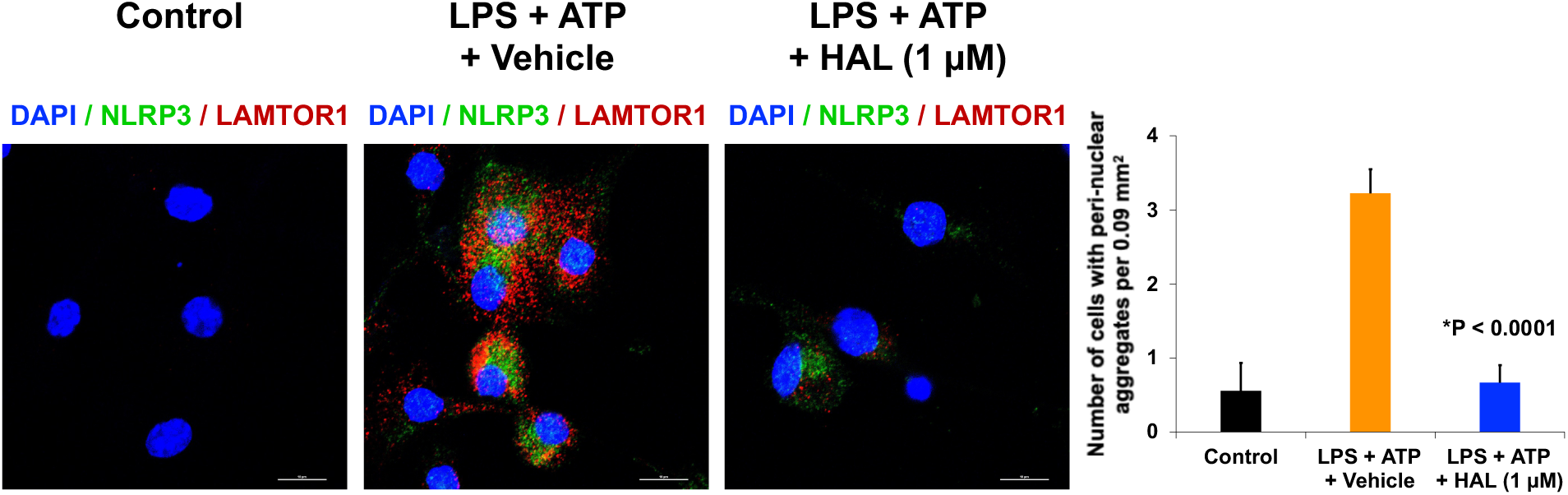
Representative immunofluorescent images show LPS + ATP induces peri-nuclear (blue, DAPI) aggregation of NLRP3 (green) and LAMTOR1 (red) in macrophages. Haloperidol (HAL, 1 µM) reduces LAMTOR1–NLRP3 aggregation. Bar graph of mean and SEM. N = 9 per group. *P < 0.0001 compared to LPS + ATP, two-tailed Student t test.

## Discussion

We identified a significant association between haloperidol use and a reduced risk of incident gout. A strength of the insurance database analysis is its large size, which constitutes a majority of all U.S. adults. Another strength is adjustment for confounders and propensity score matching, which simulates randomization and increases the validity of the conclusion. Nevertheless, as this retrospective study was not randomized, there could be residual confounding or selection bias. We also present biochemical evidence that haloperidol reduces inflammasome activation in cell culture systems. Our discovery of LAMTOR1 as a novel interacting partner of haloperidol could provide further insight into the full spectrum of this drug’s effects. Given that LAMTOR1 is a chaperone for NLRP3, haloperidol could also be used as a tool to dissect inflammasome assembly and activation.

These investigations collectively suggest a potential beneficial effect of haloperidol in forestalling gout onset. These studies also provide a rationale for performing randomized controlled trials of haloperidol for gout, which can provide insights into causality. Traditional approaches to drug development consume more than a decade and nearly $3 billion, with more than 99% of drug candidates failing (30). Our identification of this unrecognized activity of an existing FDA-approved drug could accelerate a new therapeutic approach for gout.

The tissue concentration of haloperidol in schizophrenia patients is 10 µM (31, 32). We found that lower concentrations of haloperidol (0.1–1 µM) also could reduce inflammasome activation. Therefore, doses of haloperidol lower than currently prescribed for mental disorders might be beneficial for gout. As haloperidol is a cell-permeable small molecule, another potential mode of delivery is a sustained release implant or transdermal patch. Alternatively, it could be directly injected into affected joints. Such strategies could limit potential side effects.

## DISCLOSURE

V.L.A, B.D.G., B.C.W, and S.W. are named as inventors on patent applications filed by the University of Virginia.

## Data Availability

All data produced in the present work are contained in the manuscript.

